# Combining tumor genomic and transcriptomic analysis with liquid biopsy ctDNA monitoring: analytical validation and clinical insights

**DOI:** 10.1101/2025.07.12.25331415

**Authors:** Nam HB Tran, Thien-Phuc Hoang Nguyen, Van-Anh Nguyen Hoang, Tien Anh Nguyen, Minh-Duc Nguyen, Ha-Hieu Pham, Tho Thi Le Vo, My TT Ngo, Duy Sinh Nguyen, Hoai-Nghia Nguyen, Minh-Duy Phan, Hoa Giang, Lan N Tu

## Abstract

**Background:** Comprehensive genomic profiling (CGP) is a time– and tissue-efficient method to help guide precision oncology. To enhance clinical utility of CGP, we investigated performance of a novel strategy integrating tumor DNA and mRNA profiling, together with liquid biopsy ctDNA monitoring.

**Methods:** Genomic DNA and mRNA simultaneously extracted from 604 archived tissue samples of 12 cancer types were used. Tumor DNA was subjected to targeted sequencing using a 504-gene panel with high-density probes (HDP), and shallow whole genome sequencing to profile genomic biomarkers. mRNA transcriptome profiling was performed to further capture fusion variants, and to predict tissue of origin (TOO) using our ensemble model OriCUP, an algorithm trained on 9,889 samples and independently validated on 731 samples. In a cohort of 55 metastatic lung cancer patients, longitudinal plasma ctDNA was analyzed using a hybrid tumor-informed and tumor-agnostic approach to predict progression-free survival (PFS).

**Results:** Among all biomarkers, DNA-sequencing using HDP achieved higher sensitivity than the standard panel design to identify copy number variations at chromosome-, gene– and exon-levels. The detection rate of fusion variants using DNA-sequencing alone was 20% lower than mRNA-sequencing in reference samples, while combination of both methods was essential to maximize fusion detection in clinical FFPE samples. For TOO, our OriCup model achieved prediction accuracy of 87.7% for primary tumors and 81.4% for metastatic tumors. In 55 lung cancer patients, ctDNA profiling identified additional 11.5% tumor-agnostic actionable and resistance mutations. Patients having more than 50% ctDNA decrease from baseline were classified as molecular responders, and showed significantly longer PFS than those classified as molecular non-responders (HR=9.42, 95% CI: 3.33-26.67, p<0.0001, 12-month PFS: 95.5% vs 31.7%).

**Conclusions:** Comprehensive genomic and transcriptomic profiling could reliably unveil genetic details not provided by DNA-only CGP. The integration of ctDNA detection further helped detect tumor-agnostic mutations and monitor treatment response.

## Introduction

The fast pace of biomarker discovery and increasing number of approved targeted therapies are driving the demand for comprehensive genomic profiling (CGP) in precision oncology [1]. Unlike single-gene assays or small gene panels, a comprehensive approach maximizes the chances of identifying relevant biomarkers for patient stratification, predicting drug response and monitoring resistance mechanism. Majority of current CGP utilizes formalin-fixed paraffin embedded (FFFE) tissue samples as their high tumor content allows for high sensitivity and specificity in detecting genetic alterations [2]. FFPE DNA is more commonly used than FFPE mRNA because DNA, despite damages from formalin fixation process, is generally more stable and robust compared to mRNA. However, mRNA profiling is particularly sensitive to assess novel or atypical gene fusions that DNA analysis alone may not detect [3]. Moreover, the tumor transcriptome could help predict tissue of origin (TOO) for cancer of unknown primary (CUP), which can then be combined with the genetic alteration profile to guide more effective site-specific treatment for patients with CUP [4]. Although it is evident that mRNA profiling could complement DNA information to expand therapeutic options, the current practice of running mRNA and DNA testing separately is inefficient in terms of both time and tissue utilization.

While tissue biopsies provide a direct snapshot of the molecular landscape of primary tumors, they are invasive, subject to sampling bias, and may not fully capture tumor heterogeneity or dynamic changes over time [5, 6]. Liquid biopsy (LB), conversely, are minimally invasive and serial monitoring of circulating tumor DNA (ctDNA) in LB assists real-time tumor burden assessment and hence prediction of treatment response [6]. LB-ctDNA profiling is typically performed as a separate test from FFPE profiling, resulting in less comprehensive information and ultimately higher cost for patients.

Since FFPE DNA and mRNA profiling together with LB-ctDNA detection, each offers unique and complementary information, combining them into a single affordable test could potentially unlock more treatment opportunities for cancer patients. In this study, we validated the technical performance and presented the clinical relevance of our unified approach that integrated profiling of tumor DNA using high-density probes, tumor mRNA transcriptome, and LB-ctDNA.

## Materials and Methods

### Sample and data collection

A total of 604 archived tissue samples of 12 cancer types were obtained from the Medical Genetics Institute, Viet Nam. All samples were de-identified and stored for less than 2 years. For FFPE-derived biomarkers, 241 and 375 tissue samples were used for DNA and mRNA analysis respectively. Specifically for TOO prediction, transcriptome data of 9,889 samples from TCGA database [7] and 356 samples from Robinson *et al* [8] were added to the analysis. For LB-ctDNA assessment, 147 real-world plasma samples collected from 55 stage-IV lung cancer patients were analyzed and compared with clinical records, following our published study design [9]. Detailed sample information for each analysis is listed in Table S1.

Ethics for the FFPE-derived biomarker analysis and the real-world ctDNA monitoring study were approved by the Institutional Ethics Review Board of the Medical Genetics Institute: approval 03/2024/CT-VDTYH and 03/2023/CT-VDTYH respectively. All participants provided written informed consent for the use of their anonymized genomic and clinical information.

### Sample processing

Genomic DNA and total RNA were co-isolated from FFPE by AllPrep DNA/RNA FFPE Kit (Qiagen, Germany). DNA libraries from matched FFPE and white blood cells (WBC) were prepared as described previously [10] and hybridized with probes targeting 504 gene (Integrated DNA Technologies, USA) (Table S2). The standard panel design (STD) had equal probe density in all interrogated coding regions, while the high-density probe (HDP) panel was enriched with more probes in certain chromosomes and 27 genes that often have copy number variation (CNV). Total RNA was converted to cDNA library using NEBNext® Ultra™ II Directional RNA Library Prep Kit (New England Biolabs, USA), followed by exome enrichment of 19,435 genes (xGen™ Exome Hyb Panel v2, Integrated DNA Technologies, USA). Enriched libraries were sequenced using DNBSEQ-T7 sequencer (MGI Tech, China). For shallow whole-genome sequencing (sWGS), FFPE DNA libraries were sequenced directly using DNBSEQ-G400 sequencer (MGI, China) with an average depth of 1-2X. Besides clinical samples, 14 reference samples (Table S3) were processed using the same protocol.

Plasma ctDNA analysis was performed using a hybrid tumor-informed and tumor-agnostic approach that examined both mutation and non-mutation features of ctDNA (K-4CARE, Gene Solutions) [10]. To determine limit of detection (LOD), clinical plasma samples with pre-determined tumor fraction (TF) levels of 0.5% were serially diluted to TF levels of 0.1%, 0.05%, 0.025%, and 0.01%, and analyzed in triplicates.

### DNA sequencing data analysis

Somatic variant calling, using tumor–normal mode, and germline variant calling were performed using DRAGEN™ (v4.3) as described previously [10, 11]. CNV was analyzed using DRAGEN’s somatic CNV pipeline (v4.3); gene amplification was defined when the ratio of segment mean was ≥ 2.0, while gene loss was defined when the ratio was below 0.6. In-silico tumor fraction (TF) was estimated using FACETS (v0.6.2), based on heterozygous SNPs present in both FFPE and WBC samples. The absolute copy number (ACN) was then estimated using the formula: ACN=([2*ratio – 2(1 – TF)])/(TF). For large genomic rearrangement (LGR) at exon-level, the DRAGEN Structural Variant Caller pipeline utilizing both paired-end and split-read mapping information was used. For homologous recombination deficiency (HRD) analysis, besides *BRCA* mutations, data from sWGS were processed to calculate the whole-genome instability (wGI) score as described previously [12]. Samples with a wGI score ≥ 0 were classified as positive for genomic instability. Chromosome-level structural variants (SVs) were also detected using sWGS data by calculating the log2 ratio of each target bin size relative to the median value across the genome, exclud ing sex chromosomes. A log2 ratio below 0.5 indicated a loss event, while a log2 ratio above 1.3 indicated a gain event [13].

### mRNA sequencing data analysis

For fusion detection, FASTQ files were pre-processed to remove low-quality bases before alignment to the GRCh38 reference genome. DRAGEN (v4.3) in RNA mode was used for read alignment and fusion detection. Samples were excluded from further analysis if they failed QC criteria: total input reads ≤ 30 million, median insert size < 100 bp, or coefficient of variation in median transcript coverage > 1. A fusion event between 2 genes was considered positive if the number of junction-supporting reads was ≥ 5. Fusions involving exon-skipping events within a single gene were considered positive if supported by more than 10 reads.

### Machine learning model for tissue of origin prediction

Gene-level read counts from processed BAM files were obtained using the featureCounts tool from the Subread package (v.2.0.1) [14]. Raw read counts were normalized to remove batch effects using the TMM (Trimmed Mean of M-values) method in the edgeR package (v.4.1.2) and then converted to FPKM values. In addition to in-house samples, normalized expression profiles from 32 primary tumor types (n = 9,889) were downloaded from the UCSC Xena platform [7] in FPKM format (Table S1). These samples were split into training set (8-100 samples/cancer type) and testing set (n=2,803 and 7,086 samples, respectively) to optimize machine learning (ML) model. The independent validation dataset had 731 samples consisting of 355 metastatic tumors from Robinson *et al* [8] and in-house 376 primary and metastatic tumors (Table S1).

For model training, we first established 3 gene sets: 90 genes from Ye *et al* study [15]; 2000 high-variable genes selected by a modified SIMBA method [16] that identified top variable genes to distinguish cancers in the training set with gini score ≥ 0.3; and 739 high-variable genes that were further selected from the 2000 genes above to enable classification accuracy >50% in common cancer types. Using the 3 gene sets, we then used the AutoML framework auto-sklearn (v.0.15.0), based on scikit-learn package, to train our OriCUP model with multi-class classification approach. The pipeline tested various supervised learning algorithms with hyperparameter optimization via 10-fold cross-validation. The final ensemble model was selected based on the lowest loss across validation folds, and the gene set was selected for the highest prediction performance in the validation dataset. For each sample, the top two predicted labels with the highest probabilities were reported.

The OriCUP model was compared with the deconvolution method [17] and the CUP-AI-Dx model [18]. Deconvolution was performed using FARDEEP (v.1.0.1), and normalized expression profiles from both TCGA cancer samples and GTEx normal tissues were used as input with 3 gene sets as signature markers for TOO. The CUP-AI-Dx model was downloaded from its GitHub repository (https://github.com/TheJacksonLaboratory/CUP-AI-Dx) and the normalized TPM data were adjusted to meet the input requirements.

## Statistical analysis

Survival curves were estimated by the Kaplan–Meier method, with group differences assessed by the log-rank test. Cox proportional hazard regression was used to calculate the hazard ratio (HR). The Student’s t-test was used for comparison of copy number quantification between two panel designs. All statistical analyses were conducted using GraphPad Prism v.10.3.0 with significance set at *p* < 0.05.

## Results

### Assay design

Current CGP tests using only FFPE DNA profiling had some disadvantages (Figure 1A). For MSI, TMB, and mutations (SNPs, indels), tumor-only analysis is less accurate than tumor-normal analysis; DNA-based fusion detection has lower sensitivity than mRNA-based profiling; and several biomarkers such as CNV, HRD and germline mutations require separate testing (Figure 1A). Therefore, our assay design integrated FFPE DNA-RNA and LB-ctDNA profiling to address these gaps (Figure 1B). First, DNA from paired FFPE and WBC were hybridized with a 504-gene HDP panel to determine MSI, TMB, mutations (SNPs, indels, common fusions), CNV and LGR in tumor-normal analysis mode. FFPE DNA libraries were also subjected to shallow WGS to detect CNV at chromosome level and assess genome instability status for HRD analysis. Second, FFPE mRNA-sequencing was used to enhance sensitivity for fusion detection and to predict TOO for CUP using ML model. Third, plasma cfDNA was subjected to hybrid tumor-informed and tumor-agnostic testing. The first multiplex PCR reaction utilized a fixed hotspot panel to identify tumor-agnostic mutations, including resistance mutations. The second reaction used ultra-deep sequencing to track personalized tumor-derived mutations. Together, the status and level of ctDNA helped monitor tumor burden reflecting treatment response (Figure 1B).

**Figure 1.**
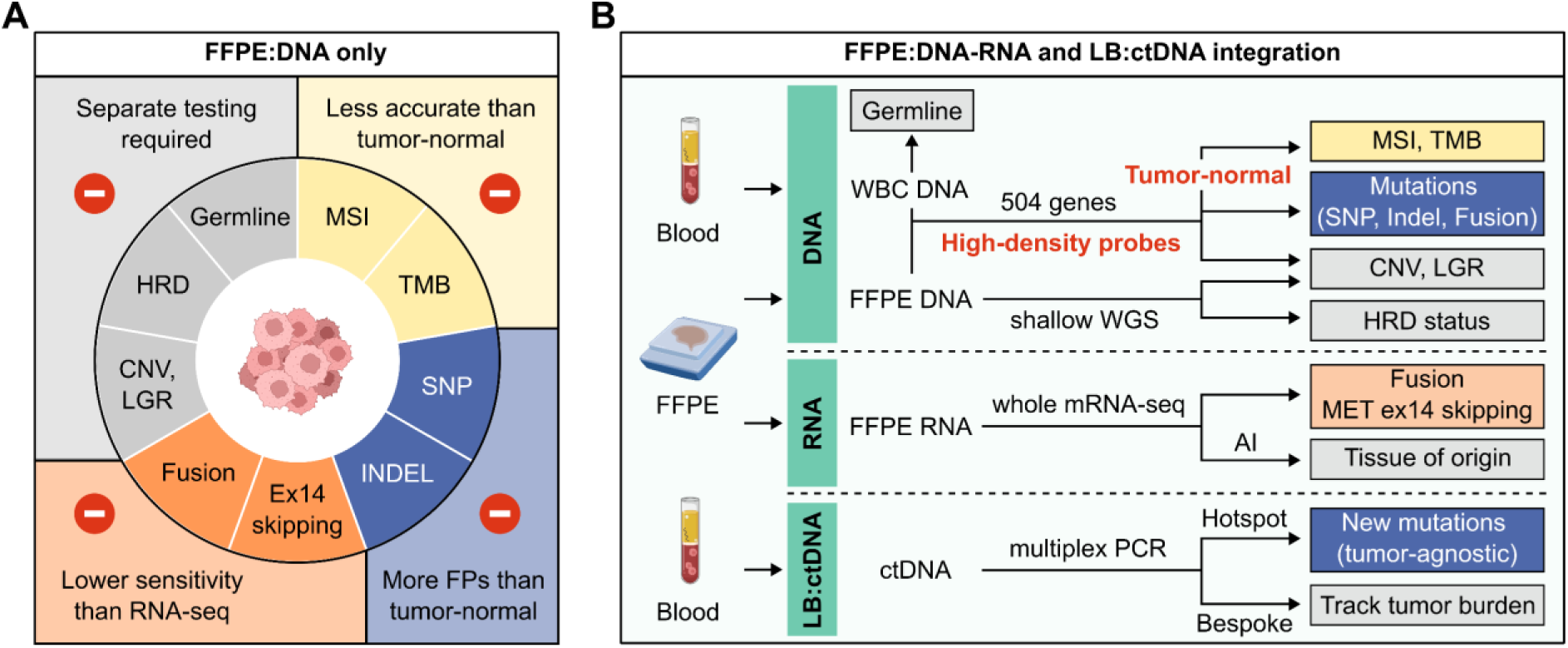
Assay design to integrate FFPE DNA-RNA and LB ctDNA profiling. **(A)** Disadvantages of CGP using only FFPE DNA to detect biomarkers. **(B)** Our design to first sequenced DNA from matched FFPE and WBC using a 504-gene panel with high-density probes and analyzed data in tumor-normal mode to determine microsatellite instability (MSI), tumor mutational burden (TMB), mutations (SNP, Indel, common fusions), copy number variation (CNV) and large genomic rearrangement (LGR). FFPE DNA was also subjected to shallow whole genome sequencing (WGS) to determine CNV, LGR and homologous recombination deficiency (HRD) status. Second, FFPE mRNA-sequencing was performed to further capture fusion variants, and also combined with machine learning (ML) to predict cancer tissue of origin. Third, plasma ctDNA was subjected to multiplex PCR reactions amplifying both cancer-specific hotspots and personalized tumor-derived mutations to identify new tumor-agnostic mutations and track tumor burden respectively.

### DNA sequencing using high-density probes to detect copy number variation

We compared performance to detect CNV at chromosome-, gene– and exon-levels between our standard (STD) and high-density probe (HDP) panels. First, SVs – DNA gain/loss at chromosome level, were analyzed in 2 glioma FFPE samples (Figure 2A). The HDP panel showed clearer signal of chromosome 1p/19q co-deletion in sample TOO289, and detected chromosome 10 loss in sample TOO293 that the STD panel failed to capture. At gene level, signals of gene amplification were also more pronounced using HDP panel compared to STD panel (Figure 2B). The deviation of absolute copy number calculated for reference samples was 0.1-0.4 copies for HDP panel, significantly lower than the variable deviation of up to 1.5 copies for STD panel (Figure 2B). For gene deletion, we detected both hetero– and homo-zygous co-deletion of *MTAP*-*CDKN2A* in reference samples using HDP panel but not STD panel (Figure 2C). In clinical lung cancer samples (n=49), the prevalence of homozygous loss of *MTAP-CDKN2A* was 12.2% (6/49) (Figure 2C); and 83.3% (5/6) of these samples also had *EGFR* co-mutation (Figure S1A). At exon level, LGRs – insertion/deletions spanning one or more exons, are challenging to capture with low probe coverage. STD panel had 81.8% sensitivity to detect LGRs of different sizes in *BRCA1/2* genes, while HDP panel achieved 100% sensitivity (Figure 2D). *In-silico* simulation of serial tumor fractions showed that LGR detection was robust at VAF of 25% (Figure S1B). Finally, the combination of CNV and *BRCA1/2* mutation detection determined HRD status in clinical samples. In 169 ovarian cancer samples, the prevalence of HRD+ was 55.0% (Figure 2E). Among HRD+ cases, the percentage of those having mutated *BRCA* alone (m*BRCA*), positive whole genome instability score alone (wGI+), and both m*BRCA* and wGI(+) together were 11.9%, 58.1%, 30.0% respectively (Figure 2E). *BRCA1* mutations were more prevalent than *BRCA2* mutations (Figure S1C).

**Figure 2.**
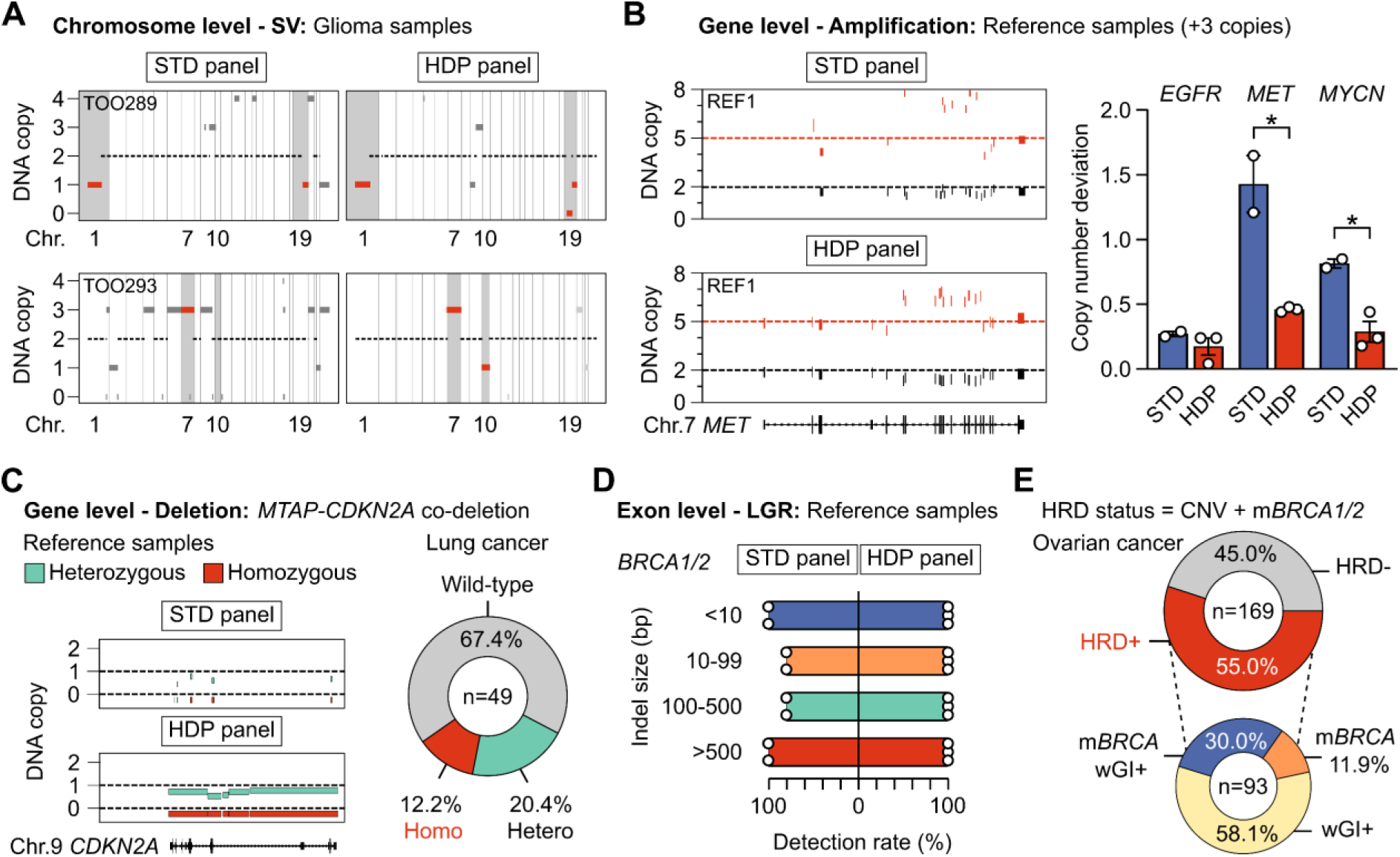
Performance of FFPE DNA sequencing using high-density probes to determine copy number variation. **(A)** Detection of structural variants (SV), or changes in copy numbers at chromosome level, in 2 FFPE glioma samples using standard (STD) panel and high-density probe (HDP) panel. **(B)** Detection of gene amplification and quantification of copy numbers in reference samples using STD and HDP panels (n=3). *p<0.05, Student’s t-test. **(C)** Detection of *MTAP*-*CDKN2A* gene loss in reference samples using STD and HDP panels. The prevalence of homozygous and heterozygous *MTAP-CDKN2A* deletion in lung cancer samples (n=49). **(D)** Detection of large genomic rearrangement (LGR), or large indel of different sizes, in *BRCA1/2* genes in reference samples using STD and HDP panels (n=20 LGRs). **(E)** Determination of homologous recombination deficiency (HRD) status in ovarian cancer samples (n=169). HRD+ was defined as either positive whole genome instability (wGI) score, determined by the presence of CNV, and/or mutated *BRCA1/2* (*m*BRCA).

### mRNA sequencing to capture fusion and predict tissue of origin

In fusion-positive reference samples, mRNA profiling achieved 100% sensitivity to capture all fusion variants, while DNA profiling detected 80% of them (Figure 3A). Some variants were not covered by our DNA probe design and hence not detected. Whole mRNA sequencing not only had broader gene coverage but also detected more fusion events than DNA sequencing, explaining its higher sensitivity. However, in real-world clinical samples, we observed high rate (35%) of FFPE samples having low RNA quality with DV200 below 30% (Figure 3B). Therefore, compared to DNA-or mRNA-profiling alone, combination of both methods was essential to maximize the sensitivity to detect actionable fusions in clinical samples (Figure 3C). In addition, novel fusion variants reported in the literature could also be identified in clinical samples for research purposes (Figure S2A). Finally, the detection of *MET* ex14 skipping variants also showed higher sensitivity using mRNA-than DNA-sequencing (Figure S2B). All performance and quality control criteria are listed in Table 1.

**Figure 3.**
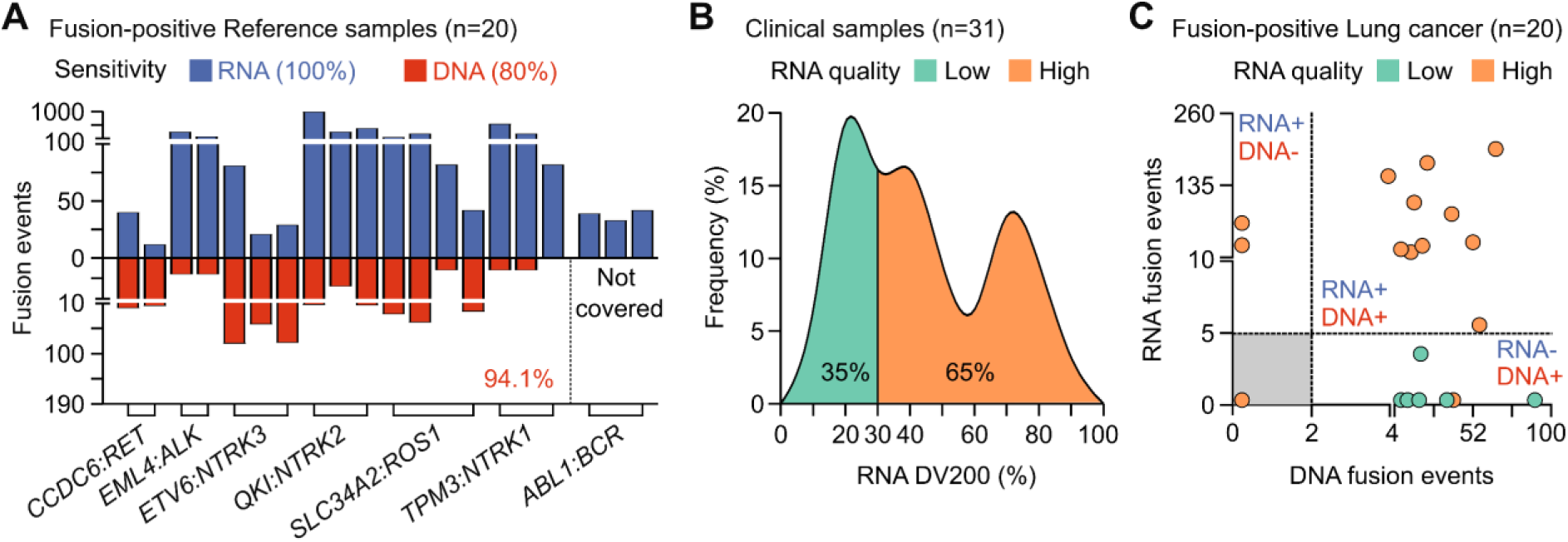
Performance of FFPE DNA and mRNA sequencing to detect fusion variants. **(A)** Comparison of 2 methods to detect fusion variants in reference samples (n=20 mutations in 10 reference samples). mRNA profiling showed more fusion events and had broader coverage of fusion genes than DNA targeted sequencing. **(B)** mRNA quality determined by DV200 values in real-world clinical samples (n=31). **(C)** Combining DNA and mRNA sequencing maximized the fusion detection rate for lung cancer FFPE samples (n=20).

**Table 1.** Analytical performance and quality control criteria of the assay.

| ANALYTICAL PERFORMANCE |  |  |  |  |
| --- | --- | --- | --- | --- |
|  |  | Sensitivity* | Specificity* | Positivity threshold |
| Immunotherapy (FFPE)<sup>1</sup> | TMB | 94% | >99% | ≥10 muts/Mb |
|  | MSI | R<sup>2</sup> correlation with PCR: 97% |  | ≥20% |
| Targeted therapy (FFPE) | Somatic variants |  |  |  |
|  | DNA-based detection<sup>1</sup> |  |  |  |
|  | SNVs | >99% | >99% | ≥5% VAF** |
|  | INDELs (<20 bp) | >99% | >99% |  |
|  | LGR (>50 bp) | >99% | >99% | ≥25% VAF |
|  | Amplification | >99% | >99% | ≥2 copy ratio |
|  | Fusion (selected genes) | 94% | >99% | ≥2 reads at breakpoint |
|  | RNA-based detection |  |  |  |
|  | Fusion (all genes) | >99% | >99% | ≥5 reads |
|  | MET exon14 skipping | >99% | >99% | ≥10 reads |
| Hereditary cancer (WBC)<sup>1</sup> | Germline variants |  |  |  |
|  | SNVs | >99% | >99% | N/A |
|  | INDELs (<20 bp) | >99% | >99% | N/A |
| HRD status (FFPE)<sup>2</sup> | Genomic instability | OPA<sup>#</sup> = 96.3% |  | wGI ≥0 |
|  | BRCA1/2 mutations | >99% | >99% | ≥5% VAF** |
| QUALITY CONTROL CRITERIA |  |  |  |  |
| Interrogated genomic regions | DNA: coding regions of 504 genes<br>RNA: whole coding regions of 19,435 genes |  |  |  |
| Tumor content | ≥20% (for HRD: ≥30%) |  |  |  |
| Median depth | FFPE: 300X and >90% sites covering at 200X<br>WBC: 200X and >90% sites covering at 100X<br>mRNA transcriptome: 30M total reads |  |  |  |

\* Sensitivity and specificity were determined using standard reference samples: Seraseq® gDNA TMB Mix, Seraseq® FFPE BRCA1/2 LGR Reference Material, Seraseq® Breast CNV Mix, Seraseq® Lung & Brain CNV Mix (SeraCare, USA); Oncospan gDNA, BRCA germline I gDNA, Pan-Cancer 6-Fusion Panel, Tru-Q0 DNA Reference Standards (Horizon, USA); EMQN RNA fusion reference samples.
\*\* Somatic SNVs and INDELs at 1% ≤ VAF < 5% were detected with 82% sensitivity and >99% specificity.

For TOO prediction, we first used mRNA transcriptome of TCGA samples as the input for ML model training and selection (Figure 4A). Three gene sets: 90 genes from Ye *et al* study [15], 2000 genes and 739 genes selected by SIMBA method, were used to distinguish 32 types of primary tumors (Figure S3A). Each individual ML pipeline together with preprocessor and feature processor was utilized for hyperparameter optimization via 10-fold cross-validation. The best ensemble model selected based on the lowest loss was Libsvm SVC for 90-gene set, and Liblinear SVC for SIMBA-2000 and SIMBA-739 gene sets (Figure 4B). The sensitivity, specificity and AUC of these models were stable across 10-fold cross-validation (Figure S3B). Among the 3 gene sets, there was no difference in the prediction accuracy using TCGA data with only primary tumors (Figure S3C). In the validation dataset that included both primary and metastatic tumors, the SIMBA-2000 gene set and its optimal Liblinear SVC model showed a slightly higher prediction accuracy than the other 2 sets and hence was selected for the OriCUP model (Figure 4C). Using the same SIMBA-2000 gene set, we also explored deconvolution method, which deconstructed the mixed signals from bulk mRNA sequencing data into individual tissue types based on signature marker genes. Performance of this method was lower than the ML model (Figure 4D). Lastly, we benchmarked the final OriCUP (2000 genes, ensemble model) with the commonly used CUP-AI-Dx (817 genes and 1D-Inception model). Although the performance to predict TOO for primary tumors was not different, the prediction accuracy for metastatic tumors using OriCUP was overall 10% higher than the CUP-AI-Dx (Figure 4E). The difference was most pronounced for lung and gastrointestinal cancers. The overall prediction accuracy of OriCUP was 87.7% for primary tumors and 81.4% for metastatic tumors.

**Figure 4.**
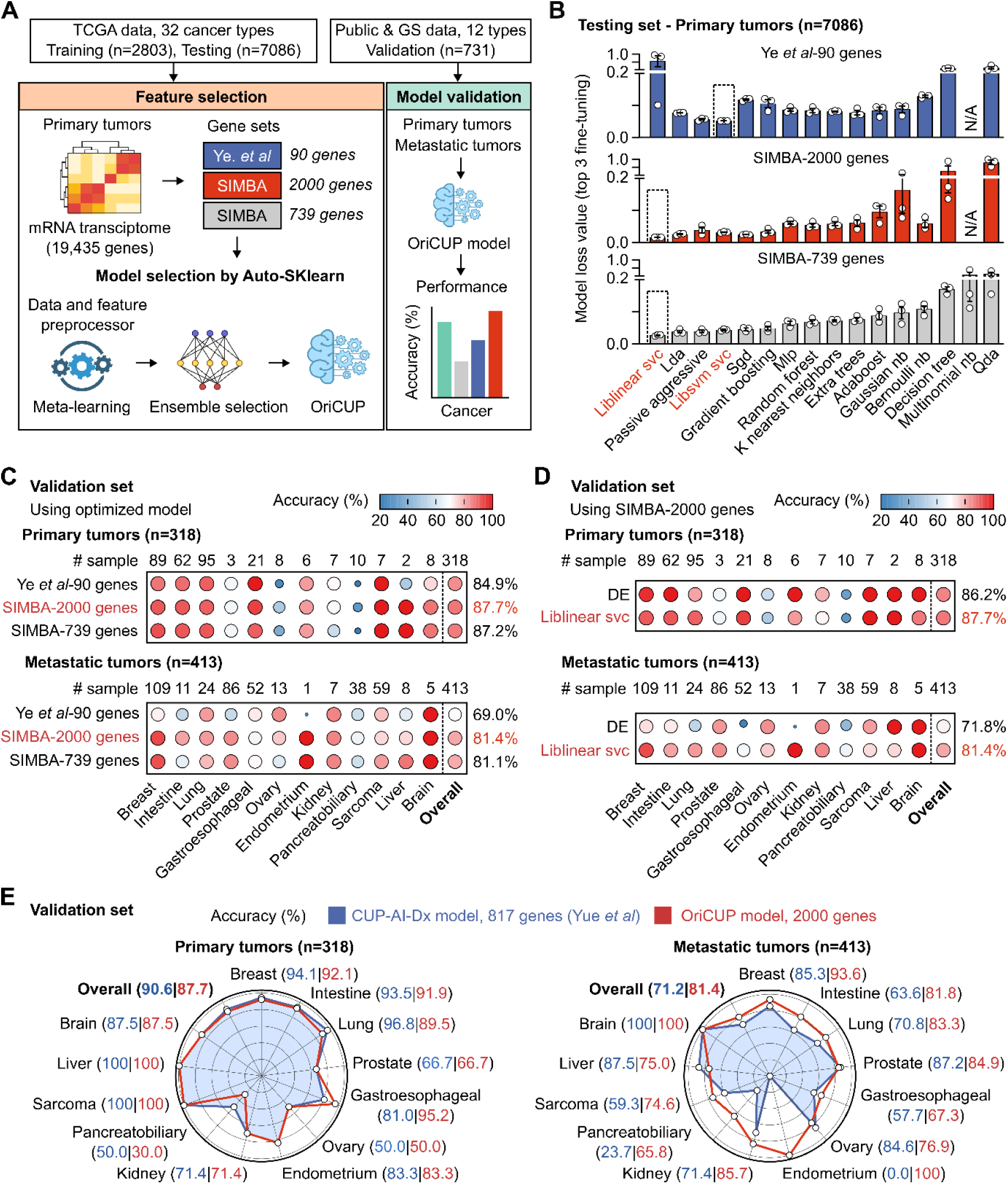
Performance of mRNA sequencing to predict cancer tissue of origin. **(A)** Workflow to develop OriCUP model to predict tissue of origin (TOO). mRNA profiles of TCGA samples (n=9,889) and the 3 gene sets were used as inputs for model training using the Auto-Sklearn framework. Performance was evaluated on an independent validation dataset (n=731) consisting of public and in-house data of Gene Solutions (GS). **(B)** Loss values of different models in the testing dataset after 10-fold cross-validation. Ensemble model having the lowest loss value was selected for each gene set. **(C)** Performance to predict TOO in the validation dataset using different gene sets and their optimized models. **(D)** Performance to predict TOO in the validation dataset using the SIMBA-2000 gene set and either its ensemble model or deconvolution (DE) method. **(E)** The final OriCUP model was benchmarked with the CUP-AI-Dx model in the validation dataset to predict TOO of both primary and metastatic tumors.

### ctDNA monitoring to identify tumor-agnostic mutations and predict disease progression

We examined the clinical significance of integrating LB-ctDNA with FFPE profiling for a small cohort of 55 stage-IV lung cancer patients. A total of 78 actionable mutations were identified in 52/55 patients, in which FFPE profiling detected 69 mutations (88.5%) in 50/52 patients, and LB-ctDNA profiling using a hotspot panel specific for lung cancer identified additional 9 tumor-agnostic mutations (11.5%), including resistance mutation *EGFR* T790M (Figure 5A). Besides actionable mutations, the total mutation profiles from FFPE were utilized to select the top 4-10 mutations for personalized ctDNA tracking. Among 227 FFPE-derived mutations selected for 55 patients, 114 mutations (50.2%) were detected in the LB of 42 patients (76.4%) (Figure 5B). The extra 14 tumor-agnostic mutations found using the hotspot panel increased the baseline ctDNA detection rate to 87.3% (48 patients) (Figure 5B). In this real-world cohort, 40 patients had serial ctDNA testing before and after treatment initiation with tyrosine kinase inhibitors or immune checkpoint inhibitors. Their clinical response up to 15 months was compared with ctDNA status. Mutation and non-mutation features including CNV and fragmentomics were combined to achieve LOD of ctDNA detection at 0.01% (Figure S4). Patients who had more than 50% ctDNA decrease from baseline were classified as molecular responders, and showed significantly longer progression-free survival (PFS) than patients with less than 50% ctDNA decrease, or molecular non-responders (HR=9.42, 95% CI: 3.33-26.67, p<0.0001); 12-month PFS were 95.5% and 31.7% respectively (Figure 5C). 84.0% (21/25) of molecular responders achieved clinical complete or partial response, while 93.3% (14/15) of molecular non-responders had confirmed progressive disease.

**Figure 5.**
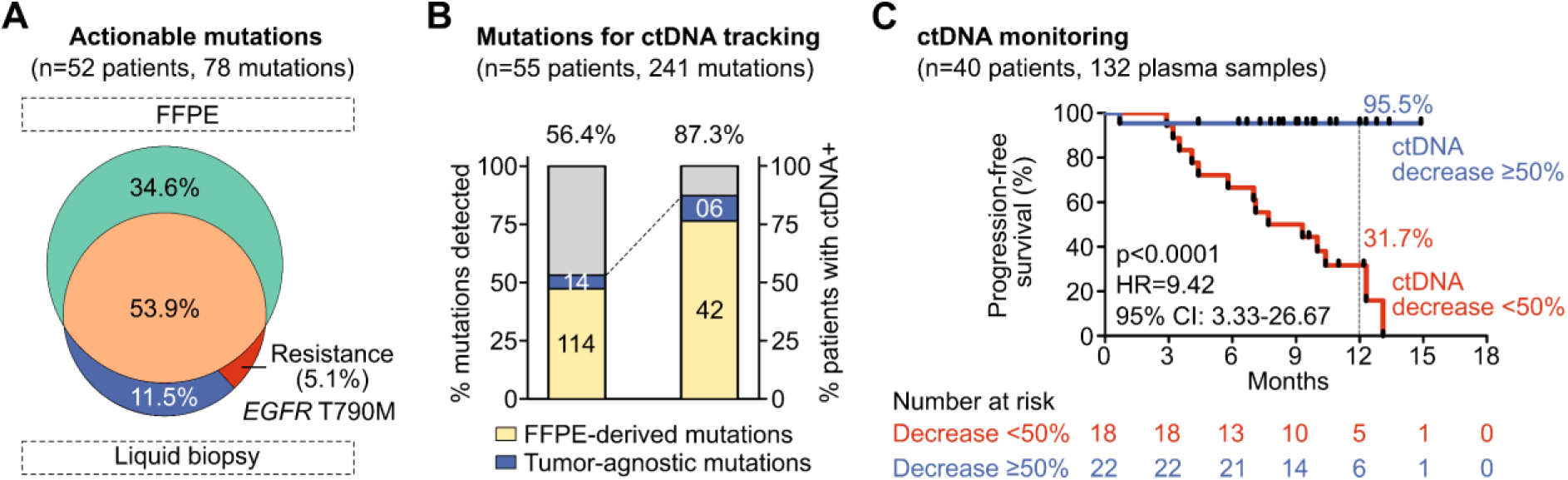
Plasma ctDNA monitoring to identify tumor-agnostic mutations and predict disease progression. **(A)** In a cohort of 55 stage-IV lung cancer patients, 78 actionable mutations were identified in 52 patients. Venn diagram showing the proportion and overlap of FFPE-derived mutations and liquid biopsy-derived mutations, including resistance mutations. **(B)** Total mutation profiles (241 mutations) were used for plasma ctDNA monitoring in 55 patients. Bar graphs showing the proportion of FFPE-derived and tumor-agnostic mutations being detected in the plasma, and the percentage of patients having positive ctDNA status before treatment. **(C)** Kaplan-Meier analysis of progression-free survival (PFS) for 40 patients as stratified by ctDNA dynamics. Patients who had more than 50% ctDNA decrease from baseline showed significantly longer PFS than patients with less than 50% ctDNA decrease (p<0.0001).

## Discussion

Current CGP testing only examines either DNA or mRNA in isolation, and uses either FFPE or LB separately. FFPE DNA remains the cornerstone of CGP assays, but the standard uniform DNA probe design had inadequate sensitivity to detect small CNVs and LGRs [19]. Therefore, increasing probe density along certain chromosomes and small genes, including part of intron regions has been demonstrated as a promising strategy [20, 21]. In this study, we showed that such HDP design indeed boosted the sensitivity to identify CNVs at both chromosome-, gene– and exon-levels and improved the accuracy to estimate copy numbers. This is particularly significant for *BRCA1/2* genes, as the prevalence of LGRs in *BRCA1/2* was reported at 2-3% in breast and ovarian patients [21–24]. Combining CNV and *BRCA1/2* mutation detection, we found that 55.0% of ovarian cancer samples were HRD+, a positive rate similar to prior studies [25, 26]. Furthermore, our HDP panel demonstrated high sensitivity to detect homozygous *MTAP*-*CDKN2A* co-deletion, an emerging therapeutic target for multiple solid tumors. The prevalence of homozygous *MTAP-CDKN2A* loss in our small lung cancer cohort was 12.2%, comparable with the reported range of 7.6%-13.4% in the literature [27–29], but a larger sample size is required for more accurate analysis. Besides, our tumor-normal analysis pairing FFPE with WBC DNA helps reduce false-positive findings as it can distinguish germline versus somatic mutations, as well as remove CHIP mutations and sequencing noise [30]. Concurrent germline testing using WBC DNA is also helpful for hereditary cancer risk assessment in high-risk patients [11, 30, 31].

Our results showed that mRNA sequencing was more sensitive to detect fusions as it covered more variants and captured more fusion events than DNA targeted sequencing, consistent with previous reports [3, 32, 33]. This is because mRNA sequencing could detect highly expressed fusion events regardless of their precise genomic breakpoints and even if the underlying DNA rearrangement is complex or cryptic [34]. However, FFPE mRNA is frequently degraded, leading to a high proportion of samples that fail quality control, a common issue reported in developing countries [35]. Therefore, combining DNA– and mRNA-sequencing could leverage the advantages of both methods to maximize patient access to fusion targeted therapies [36].

The whole mRNA transcriptome could be utilized to predict TOO for CUP, which accounts for ∼5% of metastatic cancer cases and often has poor prognosis due to limited treatment options [37]. The overall accuracy of our mRNA-based OriCUP model to predict TOO for metastatic tumors was 81.4%, higher than the mRNA-based CUP-AI-Dx model with 71.2% accuracy [18]. It was also comparable with the deep learning mRNA-based TransCUPtomics model (79.0%) [38], the DNA methylation-based BELIVE model (80.9%) [39] and the popular 90-gene qPCR method (89.2%) [15]. Previous clinical trials using mRNA-based TOO prediction alone to guide site-specific chemotherapy failed to improve survival for patients with CUP [40, 41], but the use of targeted therapies based on actionable mutation profiling alone could modestly prolonged PFS in the CUPISCO trial [42]. We speculate that identifying both TOO and actionable mutation profiles by mRNA and DNA profiling could provide stronger guidance and more treatment options for CUP patients. However, the clinical utility of this approach remains to be investigated in future randomized clinical trials.

Although many mutations detected by CGP are not therapeutically actionable, the total mutation profiles could be leveraged as “signatures” of the tumors to detect and quantify plasma ctDNA. With strong evidence from extensive research, the incorporation of LB-ctDNA monitoring has recently been proposed to complement RECIST 1.1 for more precise and earlier prediction of treatment response in metastatic cancer [43]. In addition to prognostic value, LB-ctDNA profiling could also identify resistance mutations such as *EGFR* T790M to inform therapeutic intervention. However, the required LOD of ctDNA testing and the cutoff of on-treatment ctDNA VAF reduction remains debatable. Our LB-ctDNA test with LOD of 0.01% demonstrated high sensitivity to detect pre-treatment ctDNA and showed a good correlation with clinical response, suggesting its strong potential to be used for treatment monitoring.

Although our integrated approach provides comprehensive and complex genomic data, the high cost of sequencing and analysis can be a significant barrier, limiting its widespread accessibility. The interpretation of complex genomic data requires specialized bioinformatics expertise and clinical knowledge, often leading to challenges in distinguishing between pathogenic alterations and variants of unknown significance, which can complicate clinical decision-making. To streamline the whole process and keep the assay affordable, we prioritized a small number of workflows that concurrently provided multiple biomarkers for multiple cancer types. For instance, sWGS to identify CNV could be coupled with *BRCA* mutation analysis to determine HRD status; targeted sequencing using WBC gDNA allowed better somatic mutation identification using tumor-normal analysis while also provided germline mutation detection. CGP might not be the best option for every patient, but future efforts to reduce cost and increase the number of therapeutically actionable biomarkers would help to narrow the gap between the comprehensiveness and affordability.

Although our study has not yet demonstrated clinical utility, which requires future prospective clinical studies, it showed high accuracy and robustness of the unique integrated approach of tumor genomic and transcriptomic profiling, with liquid biopsy ctDNA monitoring. This approach could support better and earlier decision-making to avoid ineffective treatment and unlock more personalized therapeutic options for cancer patients.

## Author contributions

Thien-Phuc Hoang Nguyen, Tien Anh Nguyen, Minh-Duc Nguyen, Ha-Hieu Pham, Minh-Duy Phan, Hoa Giang performed bioinformatics and statistical analysis. Nam HB Tran, Van-Anh Nguyen Hoang, Tho Thi Le Vo, My TT Ngo processed standard and clinical samples for sequencing. Duy Sinh Nguyen, Hoai-Nghia Nguyen performed clinical analysis. Thien-Phuc Hoang Nguyen and Nam HB Tran designed experiments and analyzed data. Lan N Tu conceived the idea, designed experiments, analyzed data and wrote the manuscript.

## Data availability statement

The data presented in the study are deposited in the BioProject repository, accession number PRJNA1288126.

