## Supplementary table and figure for "Combining tumor genomic and transcriptomic analysis with liquid biopsy ctDNA monitoring: analytical validation and clinical insights"

### Table S1. List of clinical samples and sequencing data used in this study

| Tissue samples | N = 960 |
|---|---|
| **DNA-sequencing** | **N = 241** |
| Glioma | 2 |
| Lung cancer | 70 |
| Ovarian cancer | 169 |
| **mRNA-sequencing** | **N = 731** |
| Fusion detection | N = 31 |
| Lung | 25 |
| Breast | 3 |
| Colorectal | 1 |
| Glioma | 2 |
| Tumor of origin (validation set)[#] | N = 731 |
| Tumor site | |
| Primary | 318 |
| Metastatic | 413 |
| Cancer type | |
| Breast | 198 |
| Lung | 119 |
| Prostate | 89 |
| Gastroesophageal | 73 |
| Intestine | 73 |
| Sarcoma | 66 |
| Pancreatobiliary | 48 |
| Ovary | 21 |
| Others | 44 |

[#]*Included data of 356 samples from Robison et al*

| Plasma samples | N = 55 |
|---|---|
| **ctDNA profiling** | **N = 55** |
| Stage IV lung cancer | N = 55 |
| Age at diagnosis, range (year) | 66 (38 – 84) |
| < 66, N | 23 |
| ≥ 66, N | 32 |
| Gender, N | |
| Female | 30 |
| Male | 25 |
| Histology subtype, N (%) | N = 55 |
| Non-small cell | 55 (100.0) |
| Adenocarcinoma | 42 (76.4) |
| Squamous cell carcinoma | 3 (5.5) |
| Unknown | 10 (18.1) |
| Small cell | 0 (0.0) |
| Treatment, N | N = 40 |
| Tyrosine kinase inhibitor | 31 |
| Immune checkpoint inhibitor | 9 |
| Clinical response*, N (%) | N = 40 |
| Complete response | 1 (2.5) |
| Partial response | 21 (52.5) |
| Stable disease | 3 (7.5) |
| Progression disease | 15 (37.5) |

** Evaluated by RECIST 1.1 criteria*

| Tumor of origin (training and testing sets): **TCGA samples, N** | | | **N = 9889** |
|---|---|---|---|
| ACC: Adrenal cancer | 79 | LUSC: Lung squamous cancer | 501 |
| BLCA: Bladder cancer | 409 | MESO: Mesothelioma | 87 |
| BRCA: Breast cancer | 1106 | OV: Ovarian cancer | 421 |
| CESC: Cervical cancer | 304 | PAAD: Pancreatic cancer | 178 |
| CHOL: Bile duct cancer | 35 | PCPG: Pheochromocytoma | 179 |
| COAD: Colon cancer | 471 | PRAD: Prostate cancer | 501 |
| DLBC: Lymphoma | 48 | READ: Rectal cancer | 166 |
| ESCA: Esophageal cancer | 184 | SARC: Sarcoma | 259 |
| GBM: Glioblastoma | 157 | SKCM: Melanoma | 103 |
| HNSC: Head & neck cancer | 520 | STAD: Stomach cancer | 412 |
| KICH: Kidney chromophobe cancer | 66 | TGCT: Testicular cancer | 150 |
| KIRC: Kidney clear cell cancer | 537 | THCA: Thyroid cancer | 505 |
| KIRP: Kidney papillary cancer | 290 | THYM: Thymoma | 120 |
| LGG: Low-grade glioma | 516 | UCEC: Endometrial cancer | 549 |
| LIHC: Liver cancer | 371 | UCS: Uterine carcinosarcoma | 57 |
| LUAD: Lung adenocarcinoma | 528 | UVM: Eye melanoma | 80 |

**Table S2. List of 504 genes**

| | | | | | | | |
|---|---|---|---|---|---|---|---|
| *ABL1* | *CDC73* | *ERCC3* | *HIST1H3A* | *LYN* | *NTHL1* | *RAC2* | *SOS1* |
| *ACVR1* | *CDH1* | *ERCC4* | *HIST1H3B* | *MALT1* | *NTRK1* | *RAD21* | *SOX17* |
| *AGO2* | *CDK12* | *ERCC5* | *HIST1H3C* | *MAP2K1* | *NTRK2* | *RAD50* | *SOX2* |
| *AKT1* | *CDK4* | *ERF* | *HIST1H3D* | *MAP2K2* | *NTRK3* | *RAD51* | *SOX9* |
| *AKT2* | *CDK6* | *ERG* | *HIST1H3E* | *MAP2K4* | *NUF2* | *RAD51B* | *SPEN* |
| *AKT3* | *CDK8* | *ERRFI1* | *HIST1H3F* | *MAP3K1* | *NUP93* | *RAD51C* | *SPOP* |
| *ALK* | *CDKN1A* | *ESR1* | *HIST1H3G* | *MAP3K13* | *OTX2* | *RAD51D* | *SPRED1* |
| *ALOX12B* | *CDKN1B* | *ETV1* | *HIST1H3H* | *MAP3K14* | *PAK1* | *RAD52* | *SRC* |
| *AMER1* | *CDKN2A* | *ETV6* | *HIST1H3I* | *MAPK1* | *PAK7* | *RAD54L* | *SRSF2* |
| *ANKRD11* | *CDKN2B* | *EZH1* | *HIST1H3J* | *MAPK3* | *PALB2* | *RAF1* | *STAG2* |
| *APC* | *CDKN2C* | *EZH2* | *HIST2H3C* | *MAPKAP1* | *PARK2* | *RARA* | *STAT3* |
| *AR* | *CEBPA* | *EZHIP* | *HIST2H3D* | *MAX* | *PARP1* | *RASA1* | *STAT5A* |
| *ARAF* | *CENPA* | *FAM175A* | *HIST3H3* | *MCL1* | *PAX5* | *RB1* | *STAT5B* |
| *ARID1A* | *CHD7* | *FAM46C* | *HLA-A* | *MDC1* | *PBRM1* | *RBM10* | *STK11* |
| *ARID1B* | *CHEK1* | *FAM58A* | *HLA-B* | *MDM2* | *PDCD1* | *RECQL* | *STK19* |
| *ARID2* | *CHEK2* | *FANCA* | *HNF1A* | *MDM4* | *PDCD1LG2* | *RECQL4* | *STK40* |
| *ARID5B* | *CIC* | *FANCC* | *HOXB13* | *MED12* | *PDGFRA* | *REL* | *SUFU* |
| *ASXL1* | *CREBBP* | *FANCD2* | *HRAS* | *MEF2B* | *PDGFRB* | *RELA* | *SUZ12* |
| *ASXL2* | *CRKL* | *FANCI* | *ICOSLG* | *MEN1* | *PDPK1* | *RET* | *SYK* |
| *ATM* | *CRLF2* | *FANCL* | *ID3* | *MET* | *PFB* | *RFWD2* | *TAP1* |
| *ATR* | *CSDE1* | *FAT1* | *IDH1* | *MGA* | *PGR* | *RHEB* | *TAP2* |
| *ATRX* | *CSF1R* | *FAT4* | *IDH2* | *MITF* | *PHOX2B* | *RHOA* | *TBX3* |
| *AURKA* | *CSF3R* | *FBXW7* | *IFNGR1* | *MLH1* | *PIK3C2G* | *RICTOR* | *TCEB1* |
| *AURKB* | *CTCF* | *FGF19* | *IGF* | *MLH3* | *PIK3C3* | *RIT1* | *TCF3* |
| *AXIN1* | *CTLA4* | *FGF3* | *IGF1* | *MLL2* | *PIK3CA* | *RNF43* | *TCF7L2* |
| *AXIN2* | *CTNNB1* | *FGF4* | *IGF1R* | *MLL3* | *PIK3CB* | *ROS1* | *TEK* |
| *AXL* | *CUL3* | *FGFR1* | *IGF2* | *MPL* | *PIK3CD* | *RPS6KA4* | *TERT* |
| *B2M* | *CXCR4* | *FGFR2* | *IKBKE* | *MRE11A* | *PIK3CG* | *RPS6KB2* | *TET1* |
| *BABAM1* | *CYLD* | *FGFR3* | *IKZF1* | *MSH2* | *PIK3R1* | *RPTOR* | *TET2* |
| *BAP1* | *CYSLTR2* | *FGFR4* | *IL10* | *MSH3* | *PIK3R2* | *RRAGC* | *TGFBR1* |
| *BARD1* | *DAXX* | *FH* | *IL7R* | *MSH6* | *PIK3R3* | *RRAS* | *TGFBR2* |
| *BBC3* | *DCUN1D1* | *FLCN* | *INHA* | *MSI1* | *PIM1* | *RRAS2* | *TMEM127* |
| *BCL10* | *DDR2* | *FLT1* | *INHBA* | *MSI2* | *PLCG2* | *RTEL1* | *TMPRSS2* |
| *BCL2* | *DDX31* | *FLT3* | *INPP4A* | *MST1* | *PLK2* | *RUNX1* | *TNFAIP3* |
| *BCL2L1* | *DICER1* | *FLT4* | *INPP4B* | *MST1R* | *PMAIP1* | *RXRA* | *TNFRSF14* |
| *BCL2L11* | *DIS3* | *FOXA1* | *INPPL1* | *MTOR* | *PMS1* | *RYBP* | *TOP1* |
| *BCL6* | *DNAJB1* | *FOXL2* | *INSR* | *MUTYH* | *PMS2* | *SDHA* | *TP53* |
| *BCOR* | *DNMT1* | *FOXO1* | *IRF4* | *MYB* | *PNRC1* | *SDHAF2* | *TP53BP1* |
| *BIRC3* | *DNMT3A* | *FOXP1* | *IRS1* | *MYBL1* | *POLD1* | *SDHB* | *TP63* |
| *BLM* | *DNMT3B* | *FUB1* | *IRS2* | *MYC* | *POLE* | *SDHC* | *TRAF2* |
| *BMPR1A* | *DOT1L* | *FUBP1* | *JAK1* | *MYCL1* | *PPARG* | *SDHD* | *TRAF7* |
| *BRAF* | *DROSHA* | *FYN* | *JAK2* | *MYCN* | *PPM1D* | *SESN1* | *TRRAP* |
| *BRCA1* | *DUSP4* | *GABRG1* | *JAK3* | *MYD88* | *PPP2R1A* | *SESN2* | *TSC1* |
| *BRCA2* | *E2F3* | *GATA1* | *JUN* | *MYOD1* | *PPP2R2A* | *SESN3* | *TSC2* |
| *BRD4* | *EED* | *GATA2* | *KBTBD4* | *NBN* | *PPP4R2* | *SETD2* | *TSHR* |

**Table S2. (cont.)**

| | | | | | | | |
|---|---|---|---|---|---|---|---|
| *BRIP1* | *EGF* | *GATA3* | *KDM5A* | *NCOA3* | *PPP6C* | *SETD8* | *U2AF1* |
| *BTK* | *EGFL7* | *GFI1* | *KDM5C* | *NCOR1* | *PRDM1* | *SF3B1* | *UPF1* |
| *CALR* | *EGFR* | *GFI1B* | *KDM6A* | *NCOR2* | *PRDM14* | *SH2B3* | *VEGFA* |
| *CAMTA1* | *EIF1AX* | *GLI1* | *KDR* | *NECTIN4* | *PREX2* | *SH2D1A* | *VHL* |
| *CARD11* | *EIF4A2* | *GLI2* | *KEAP1* | *NEGR1* | *PRKAR1A* | *SHOC2* | *VTCN1* |
| *CARM1* | *EIF4E* | *GNA11* | *KIT* | *NF1* | *PRKCA* | *SHQ1* | *WHSC1* |
| *CASP8* | *ELF3* | *GNAQ* | *KLF4* | *NF2* | *PRKCI* | *SLX4* | *WHSC1L1* |
| *CBFB* | *EP300* | *GNAS* | *KMT2A* | *NFE2L2* | *PRKD1* | *SMAD2* | *WT1* |
| *CBL* | *EPAS1* | *GPS2* | *KMT2B* | *NFKBIA* | *PTCH1* | *SMAD3* | *WWTR1* |
| *CCND1* | *EPCAM* | *GREM1* | *KMT2C* | *NKX2-1* | *PTEN* | *SMAD4* | *XIAP* |
| *CCND2* | *EPHA3* | *GRIN2A* | *KMT2D* | *NKX3-1* | *PTP4A1* | *SMARCA4* | *XPO1* |
| *CCND3* | *EPHA5* | *GSK3B* | *KNSTRN* | *NOTCH1* | *PTPN11* | *SMARCB1* | *XRCC2* |
| *CCNE1* | *EPHA7* | *H3F3A* | *KRAS* | *NOTCH2* | *PTPRD* | *SMARCD1* | *YAP1* |
| *CD274* | *EPHB1* | *H3F3B* | *LATS1* | *NOTCH3* | *PTPRS* | *SMARCE1* | *YES1* |
| *CD276* | *ERBB2* | *H3F3C* | *LATS2* | *NOTCH4* | *PTPRT* | *SMO* | *ZFHX3* |
| *CD79A* | *ERBB3* | *HGF* | *LDB1* | *NPM1* | *PVT1* | *SMYD3* | *ZFTA* |
| *CD79B* | *ERBB4* | *HIST1H1C* | *LMO1* | *NRAS* | *RAB35* | *SNCAIP* | *ZMYM3* |
| *CDC42* | *ERCC2* | *HIST1H2BD* | *LRP1B* | *NSD1* | *RAC1* | *SOCS1* | *ZNF521* |

**Table S3. List of reference samples used in this study**

| Sample | Manufacturer | Reference code | Validated markers |
|---|---|---|---|
| Seraseq® Lung & Brain CNV Mix, + 3 copies | Seracare (USA) | 0710-0414 | DNA gene amplification |
| *MTAP, CDKN2A* co-loss (CN=0) reference standard | Cobioer (China) | CBP40132 | DNA gene deletion |
| *MTAP, CDKN2A* co-loss (CN=1) reference standard | Cobioer (China) | CBP40150 | DNA gene deletion |
| Seraseq® gDNA *BRCA1/2* LGR Inherited Mutation Mix | Seracare (USA) | 0730-0568 | Large genomic rearrangements |
| EMQN01 | EMQN | | Fusion |
| EMQN02 | EMQN | | Fusion |
| EMQN03 | EMQN | | Fusion |
| EMQN04 | EMQN | | Fusion |
| EMQN05 | EMQN | | Fusion |
| EMQN06 | EMQN | | Fusion |
| EMQN07 | EMQN | | Fusion |
| EMQN08 | EMQN | | Fusion |
| EMQN09 | EMQN | | Fusion |
| EMQN10 | EMQN | | Fusion |

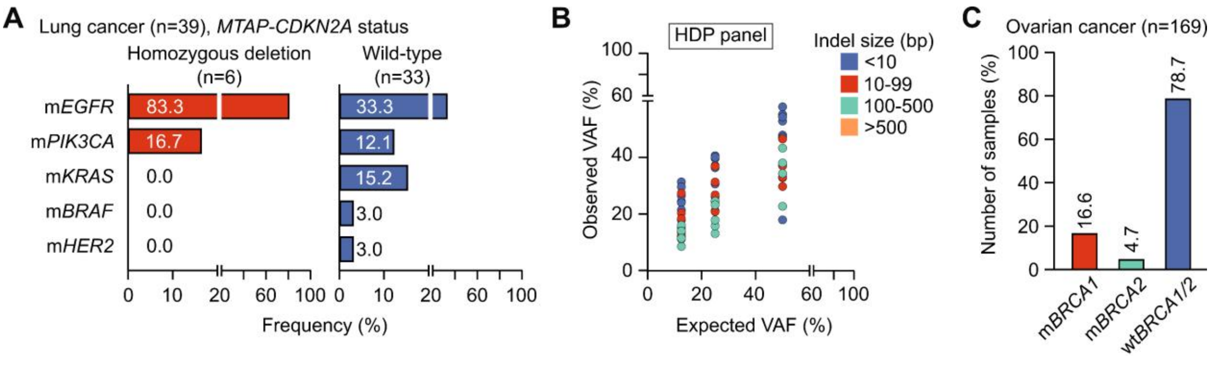


**Figure S1. Performance of FFPE DNA sequencing using high-density probes to determine copy number variation. (A)** Frequency of homozygous *MTAP-CDKN2A* deletion in lung cancer samples having other actionable mutations (n=39). **(B)** *In-silico* simulation of different VAFs for detection of large genomic rearrangement (LGR) in *BRCA1/2* genes. **(C)** Percentage of mutated *BRCA1/2* (m*BRCA*) and wild-type *BRCA1/2* (wt*BRCA*) in ovarian cancer samples (n=169).

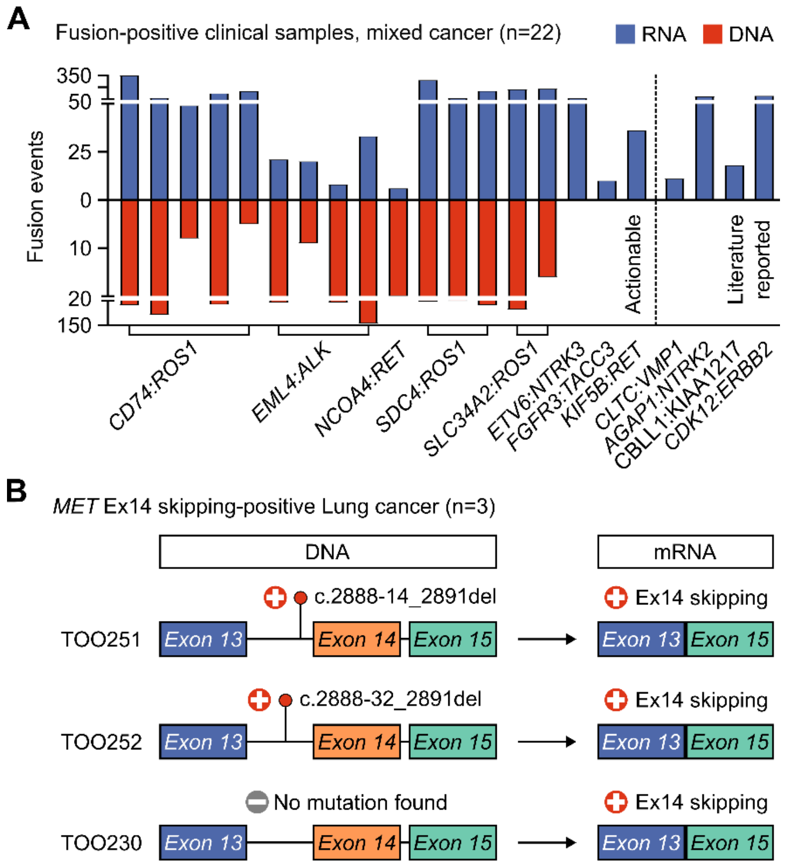


**Figure S2. Performance of FFPE DNA and mRNA sequencing to detect fusion. (A)** In fusion-positive clinical samples, mRNA profiling captured more fusion events and had broader coverage of fusion genes and partners than DNA profiling. **(B)** mRNA profiling could detect MET Ex14 skipping when no DNA mutation was identified in lung cancer samples.

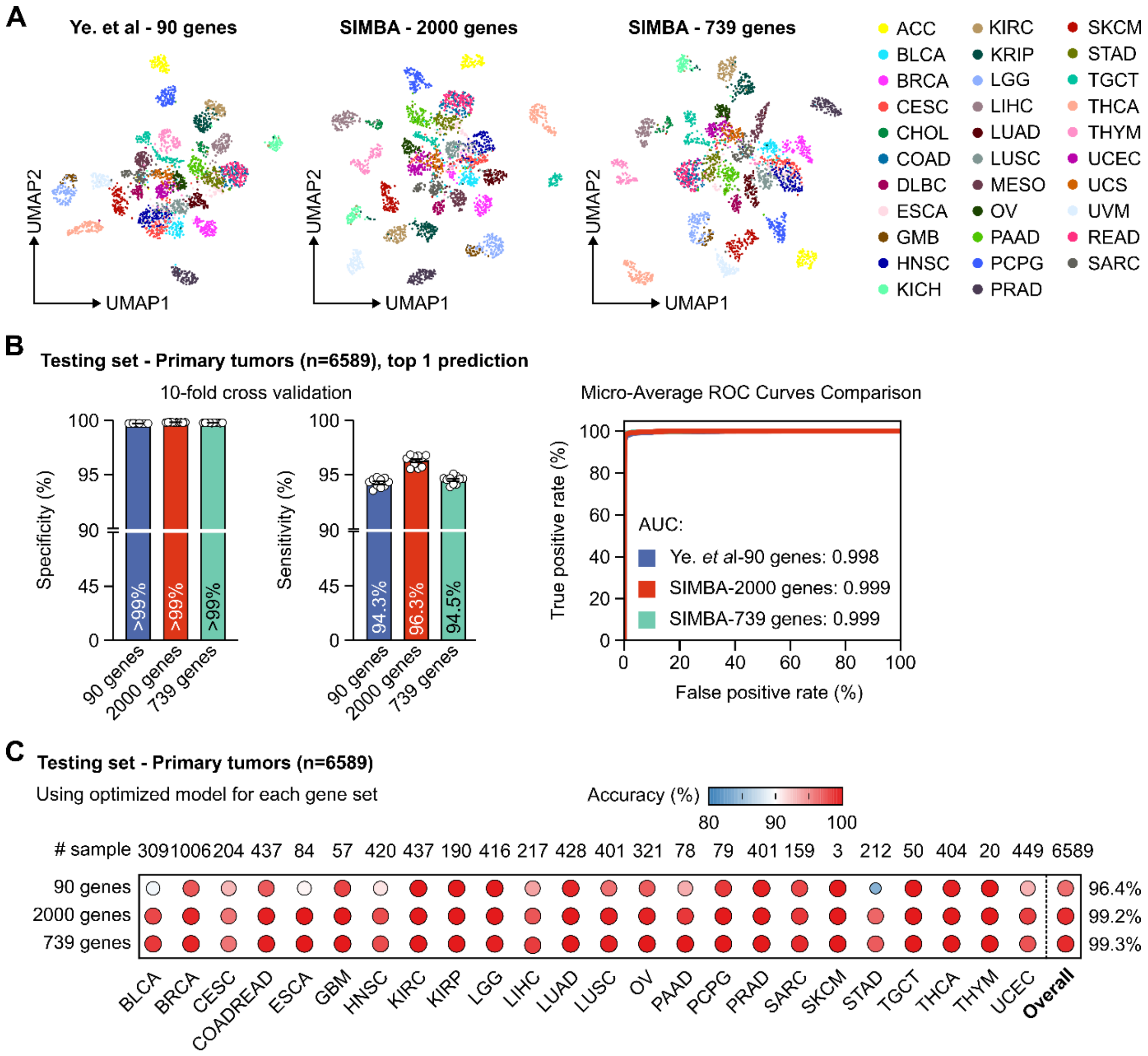


**Figure S3. Performance of mRNA sequencing to predict cancer tissue of origin. (A)** Two-dimensional Uniform Manifold Approximation and Projection for Dimension Reduction (UMAP) using 3 gene sets applied in 32 cancer types in the training dataset revealed distinct clusters corresponding to different cancer types (n=2803). **(B)** Sensitivity and specificity of optimized ensemble models in the testing dataset (n=6589) after 10-fold cross-validation across 3 gene sets. The micro-average Receiver Operating Characteristic (ROC) curves from the best cross-validation fold demonstrated stable sensitivity, specificity, and robust discriminative performance in all gene sets. **(C)** Performance to predict TOO was not different among the 3 optimized ensemble models and corresponding gene sets in the testing dataset (n=6589).

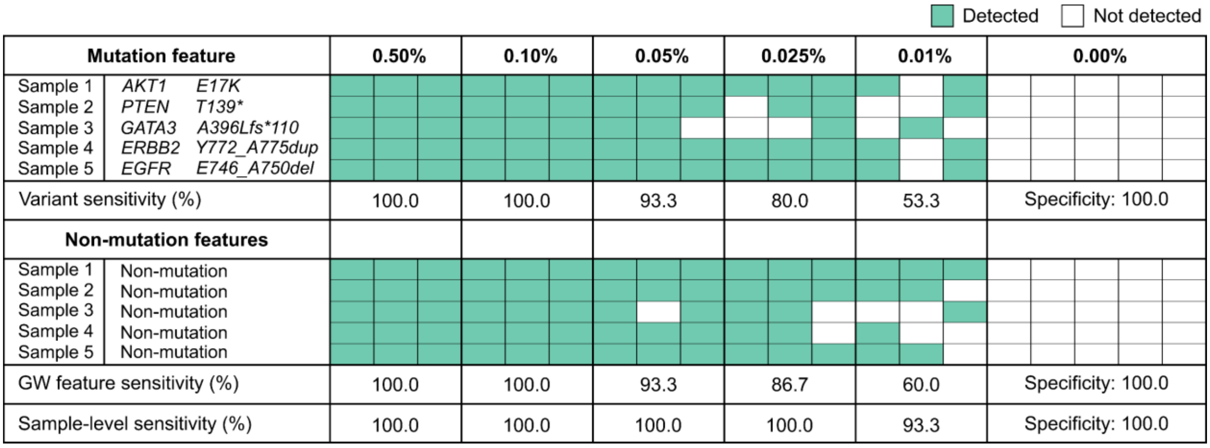


**Figure S4. Limit of detection for plasma ctDNA using combined mutation and non-mutation features**. Clinical samples were serially diluted to different levels of tumor fractions. Sensitivity to detect mutations and non-mutation genome-wide (GW) features were shown. When both mutation and non-mutation features were combined, limit of detection at 90% confidence (LOD90) was determined at 0.01%.
